# Predicting Levels of Anemia among Adolescents in Ethiopia Using homogeneous ensemble Machine Learning algorithm

**DOI:** 10.1101/2025.02.04.25321651

**Authors:** Misganaw Ketema Ayele, Eyob Teshager Enyew

## Abstract

Anemia significantly impacts adolescent girls’ health and quality of life in Ethiopia. Effective interventions require identifying key risk factors and predicting anemia severity. While traditional studies primarily use statistical methods, this research aims to leverage machine learning models to predict anemia risk and analyze contributing socio-economic, environmental, and cultural factors.

We applied machine learning models, including Random Forest, Extra Trees, CatBoost, XGBoost, and AdaBoost, to predict anemia severity using features from the Ethiopian Demographic and Health Survey (EDHS). Performance was evaluated using accuracy, ROC AUC, precision, recall, and F1-score, with feature importance analysis to identify key anemia risk factors. Random Forest and Extra Trees outperformed others, achieving accuracy rates of 82.51% and 82.41% and ROC AUC scores of 94.87% and 94.48%, respectively. CatBoost showed competitive performance (80.99% accuracy, 93.08% ROC AUC). XGBoost and AdaBoost were less effective. Key risk factors included region, education, wealth index, household size, and altitude.

Random Forest and Extra Trees are effective for predicting anemia severity and identifying key socio-economic and environmental risk factors. Interventions focusing on education, healthcare access, and nutrition are vital to reducing anemia prevalence among adolescent girls in Ethiopia. Future work should refine models and expand datasets for improved public health outcomes.

## Background

Anemia is a global public health problem that significantly impacts morbidity and mortality rates, especially among women and children [1]. According to the World Health Organization (WHO), anemia affects nearly 1.8 billion people worldwide, with a disproportionate burden on low- and middle-income countries like Ethiopia [2]. Anemia is characterized by a reduction in the number of red blood cells or hemoglobin concentration, which impairs the body’s ability to transport oxygen efficiently . It can lead to fatigue, weakness, reduced cognitive and physical performance, and, in severe cases, it can result in death if left untreated [3].

Among vulnerable groups, women of reproductive age, including adolescents, are particularly susceptible to anemia due to various biological and social factors [2], [4]. Adolescence is a critical period of physical growth and development, where nutritional needs are heightened [3]. Females in this age group are at risk of anemia due to menstruation, inadequate dietary intake, parasitic infections, rapid growth and physical changes, high iron requirements, high rate of infection and worm infestation, as well as the early marriage and adolescent pregnancy and socio-economic factors that affect access to healthcare and nutrition [4], [5].

In Ethiopia, anemia remains a pressing issue, with high prevalence rates reported among children [6], pregnant women [7], and women of reproductive age [8]. However, adolescent females (aged 15–19 years) are often overlooked, despite being a crucial demographic [9]. Addressing anemia in this group is critical for improving long-term health outcomes, as anemia during adolescence can lead to delayed growth, impaired cognitive development, and poor reproductive [1].

Existing studies [10][11][12][13][14][15] in Ethiopia have primarily used statistical analysis methods, such as bivariate and multivariate logistic regression methods, to identify risk factors associated with anemia. These methods are effective in understanding relationships between variables but have limitations in predictive accuracy. They often fail to capture the complex interactions between multiple risk factors and do not provide a clear understanding of the severity of anemia. Moreover, the studies typically categorize anemia as a binary condition (anemic vs. non-anemic), which does not reflect the reality that anemia can vary in severity and thus requires different levels of treatment and intervention. They have not generate rule that Providing data-driven recommendations to policymakers and healthcare providers on how to better allocate resources for anemia prevention and treatment. Leverage the predictive power of machine learning to offer more precise and actionable insights for early detection and intervention based on anemia severity.

There is also a growing body of machine learning research in health prediction, but much of this work has focused on other vulnerable groups, such as pregnant women [16][17][18] or children[19][20][21], and lacks a focus on adolescents. Only, the study [22] use a machine learning to predict anemia among young girls, but continue to use the binary classification approach to anemia, ignoring the distinct severity levels that have important implications for healthcare decisions.

## Related work

Several studies have examined the prevalence and determinants of anemia among adolescents [10][11][12][13][14][15]. They have used cross-sectional statistical methods like bivariate and multivariate logistic regression methods, to identify risk factors associated with anemia among adolescent females. By using these method, they investigated risk factor of anemia among adolescent are place of residence, religion, marital status, contraceptive method, number of household, educational status, occupation status and they showed that the prevalence of anima is ranged from 21% to 27% among adolescent female in Ethiopia.

The model [10][11][12][13][14][15] they used is not predictive mode, generate rules that allow the development of evidence-based strategies and policies toward preventing and/or reducing anemia of adolescent female in Ethiopia, and have limited capacity to discover new and unanticipated patterns that are hidden in data and identify the cause-and-effect relationships. Because the method the used (cross-sectional statistical methods) usually have limited capacity to discover new and unanticipated patterns and identify cause and effect relationships that are hidden in data [7]. In contrast, machine learning algorithms have demonstrated their effectiveness and efficiency in accurately predicting level of anemia among pregnant women [16][17][18] and children [19][20][21]. There is also a machine learning studies that predicts anemia among young girls in Ethiopia using Decision tree, Random forest, Extreme gradient boost, Light gradient boosting machine, Support vector machine, Logistic regression, K-nearest Neighbor, and Gaussian Naïve Bayes. Among these algorithm random forest classifier outperformed in predicting anemia among adolescent girls with an AUC value of 82%. But this studies typically categorize anemia as a binary condition (anemic vs. non-anemic), which does not reflect the reality that anemia can vary in severity and thus requires different levels of treatment and intervention.

Therefore, this research is driven by the desire to address these limitations by creating a predictive model, identifying factors that contribute to level of anemia among adolescents girls, and generate rules to support the development of evidence-based strategies, policies, and interventions aimed at preventing, managing, and reducing adolescent girls mortality in Ethiopia.

## Materials and Methods

### Materials and methods

The methodological flow chart presented in figure 1 illustrates the process followed in this study to build a predictive model for Anemia_level, identify risk factors, and extract relevant rules.

**Figure 1.**
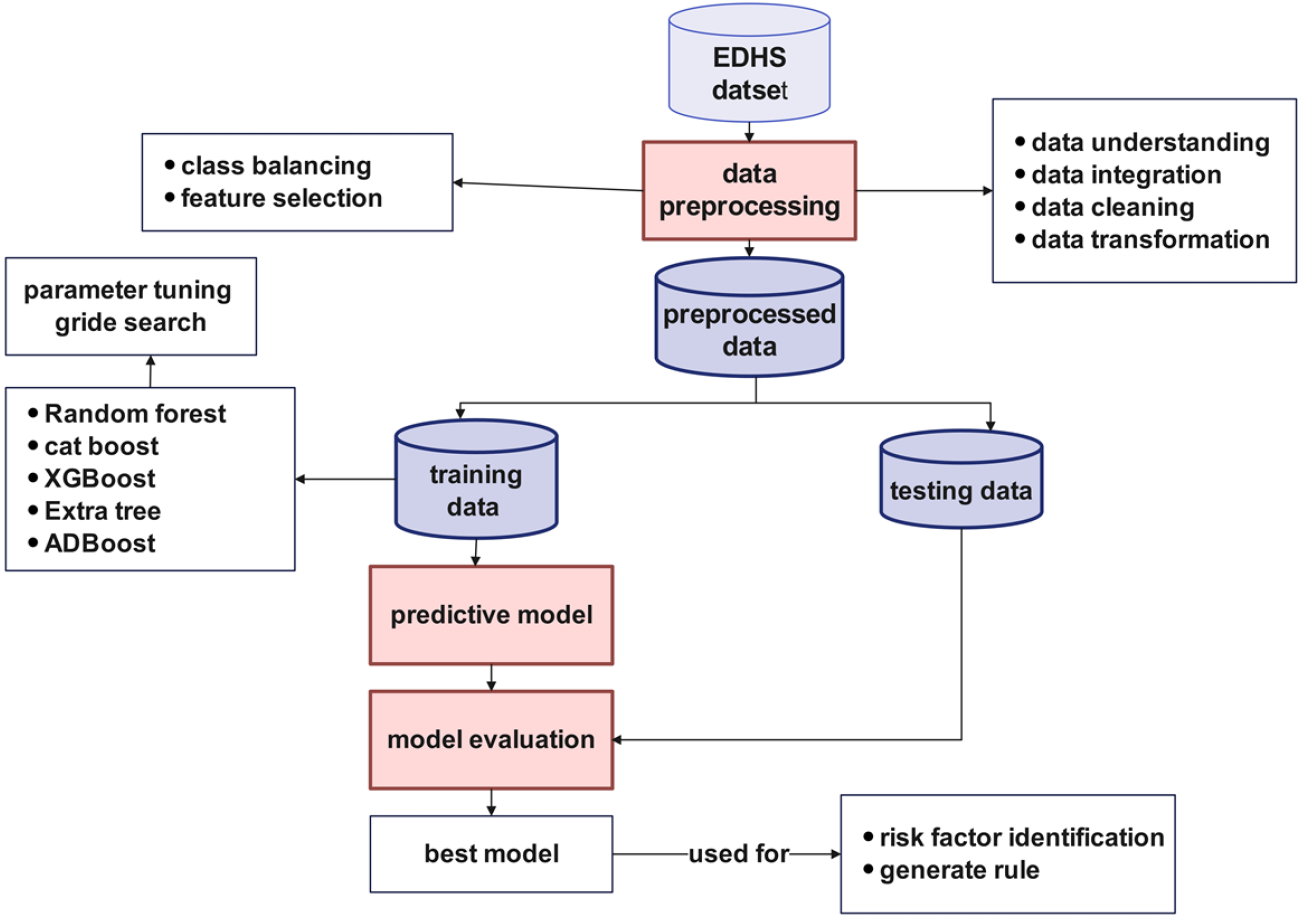
Proposed model development workflow

### Data Source

The dataset for this study is sourced from the Ethiopian Demographic and Health Survey (EDH 2011 and 2016), a nationally representative survey conducted by the Central Statistical Agency of Ethiopia in collaboration with international organizations, which is freely available online http://dhsprogram.com.. The target population for the study consists of adolescents aged 15–19 years, with a focus on predicting and identifying the factors contributing to anemia severity within this group. Key variables include region, educational status, religion, family size, sex of household head, wealth index, source of drinking water, type of toilet facility, altitude, occupation, contraceptive method, currently pregnant, currently breastfeeding, distance to health facility, marital status, media exposure, terminated pregnancy and Births in last five years. The outcome variable in this study is anemia levels, categorized into four levels: Normal, Mild, Moderate, and Severe, based on the hemoglobin thresholds defined by the World Health Organization (WHO). These variables are analyzed to predict and identify potential risk factors associated with varying levels of anemia severity in Ethiopian adolescents.

## Data preprocessing

In our research on predicting anemia levels among adolescents in Ethiopia using homogenous machine learning algorithm, we begin with **data cleaning** to handle missing values and remove duplicates. Missing values are imputed using the mode, and duplicates are removed to avoid bias. **Data transformation** follows, where categorical features are encoded into numerical values, and numerical variables are standardized to ensure fair model input. We perform **outlier detection** using methods like Z-scores to identify and address extreme values that could distort predictions.

Next, **feature selection** is carried out to identify the most relevant variables (features) from a dataset that contribute significantly to the predictive model’s accuracy, using Hybrid Methods techniques like correlation analysis and Recursive Feature Elimination (RFE), Chi-Squared Test and Sequential Feature Selection, feature importance and Recursive Feature Elimination. Among these method feature importance and Recursive Feature Elimination outperform than other. So for farther use we have used feature importance and RFE feature selection method. Irrelevant or redundant features are removed to improve model efficiency. **Class balancing** is applied to address imbalanced class distributions, ensuring equal representation for all anemia severity levels. Techniques like SMOTE are used to over-sample minority classes, preventing the model from being biased. These preprocessing steps ensure that the data is well-prepared for accurate and reliable machine learning predictions of anemia severity.

After completing all the necessary data preprocessing tasks, a total of 15952 instances with 19 features were included for further analysis and the development of the prediction model. Subsequently, the dataset was divided into training and testing datasets, following an 70/20% ratio.

## Mode development

The development of a predictive model for anemia severity in adolescents in Ethiopia leverages ensemble machine learning techniques, which combine multiple models to improve prediction accuracy and robustness. Ensemble methods, such as Random Forest, Extra Tree, Adaboost, XGBoost , and cat boost, are used for this task because they can handle complex data patterns, improve generalization, and reduce overfitting. In ensemble learning, individual base models (often decision trees) are trained independently, and their predictions are aggregated to produce a more reliable final output. Random Forest, which creates a collection of decision trees through bagging (bootstrap aggregation), is particularly effective in handling large datasets and noisy features while preventing overfitting. Similarly, Adabboost, Gradient Boosting and its variants like XGBoost and CatBoost use boosting techniques, which sequentially train models, with each new model correcting the errors made by the previous one. This iterative process increases the accuracy of the predictions by focusing on the misclassified samples.

During model training, the dataset is divided into training and testing sets, typically using an 70-30 split. To prevent model bias towards dominant classes, class balancing techniques, such as SMOTE, and feature selection are applied to handle the potential imbalance in the severity levels of anemia. The ensemble models are then trained on the balanced dataset, and hyperparameters are optimized through techniques like grid search, ensuring the best possible model performance.

After training, the model’s effectiveness is evaluated using a variety of metrics, including accuracy, precision, recall, F1-score, and ROC-AUC, to measure how well it predicts different levels of anemia severity. Cross-validation is used to assess model stability and ensure that the model generalizes well to unseen data.

Through the use of ensemble machine learning techniques, the final predictive model is not only more accurate and robust but also capable of handling the complexities and variabilities present in the dataset. The ensemble approach ensures that the model can make more reliable predictions regarding anemia severity, ultimately leading to better-informed healthcare decisions and more effective interventions for adolescents in Ethiopia.

## Model Performance

The evaluation of different homogenous ensemble machine learning models reveals varied performance across key metrics such as accuracy, ROC AUC, precision, recall, and F1-score as shown in figure 2 and 3. Random Forest and Extra Trees emerge as the top-performing models, showcasing a strong balance between accuracy and ROC AUC, with values of 82.51% and 94.87% for Random Forest, and 82.41% and 94.48% for Extra Trees, respectively. These results highlight their reliability and effectiveness in distinguishing between different classes.

**Figure 2.**
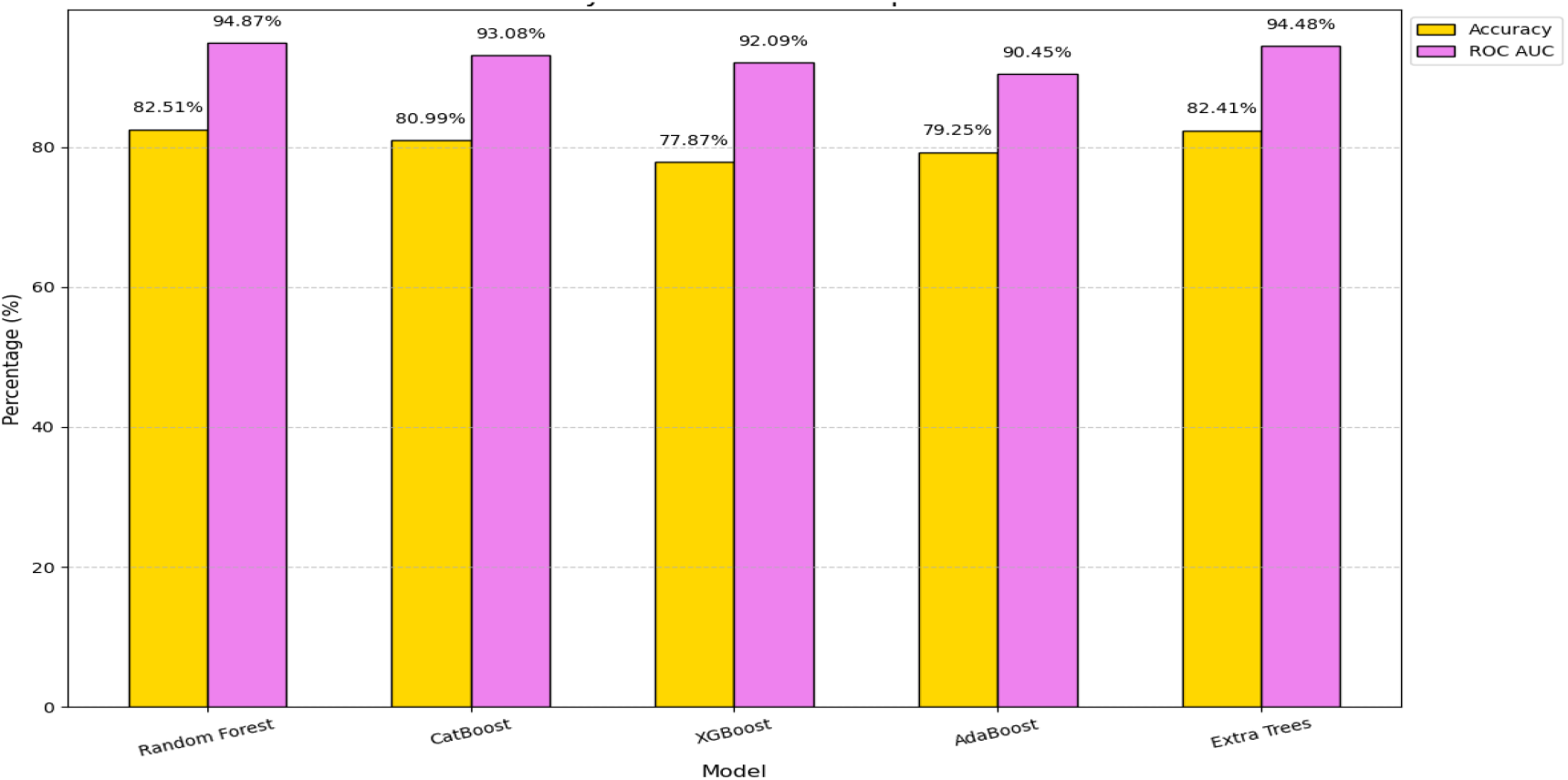
accuracy and ROC AUC comparison

**Figure 3.**
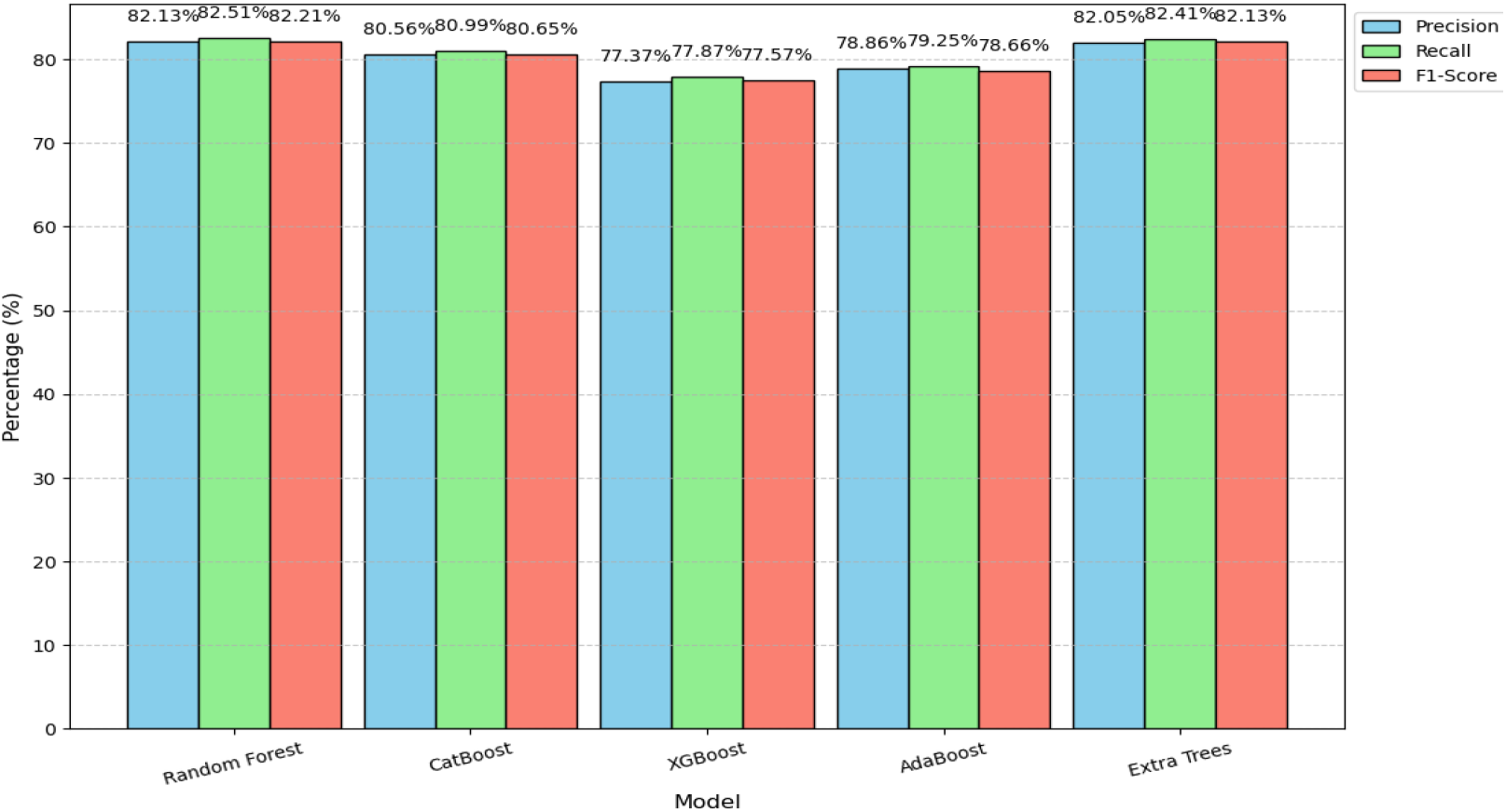
precision, recall and F1-score comparison

CatBoost also demonstrates robust performance, with a competitive ROC AUC of 93.08% and an accuracy of 80.99%, making it a strong contender despite slightly trailing Random Forest and Extra Trees. On the other hand, XGBoost and AdaBoost show lower accuracy (77.87% and 79.25%, respectively) and ROC AUC (92.09% and 90.45%, respectively), indicating they might struggle more in generalization compared to the leading models.

Precision, recall, and F1-scores are relatively consistent across all models, falling in the range of 77% to 82%, which suggests that most models handle class imbalances effectively. However, Random Forest and Extra Trees exhibit a slight edge in these metrics, reinforcing their overall superior performance. While XGBoost and AdaBoost perform adequately, they are less reliable than the other models for this task. In summary, Random Forest and Extra Trees stand out as the most suitable models for the given classification problem due to their high accuracy, excellent ROC AUC scores, and balanced performance across other metrics. CatBoost provides an alternative with solid performance, while XGBoost and AdaBoost, though competent, may require further optimization to match the top models.

### Risk factor identification

The risk factors for anemia among adolescent girls in Ethiopia reveals several key socio-economic, environmental, and cultural determinants as shown in the figure 4 below. **Region** emerged as the most important factor, indicating that geographic location significantly impacts anemia risk. This is likely due to disparities in access to nutrition, healthcare, and living conditions across different regions. For instance, girls in rural or remote areas may face higher risks due to limited access to iron-rich foods and healthcare services. Similarly, the **altitude** factor suggests that living at higher altitudes can affect iron absorption and red blood cell production, which might increase the likelihood of anemia, particularly if access to diverse food sources is limited in these regions.

**Figure 4.**
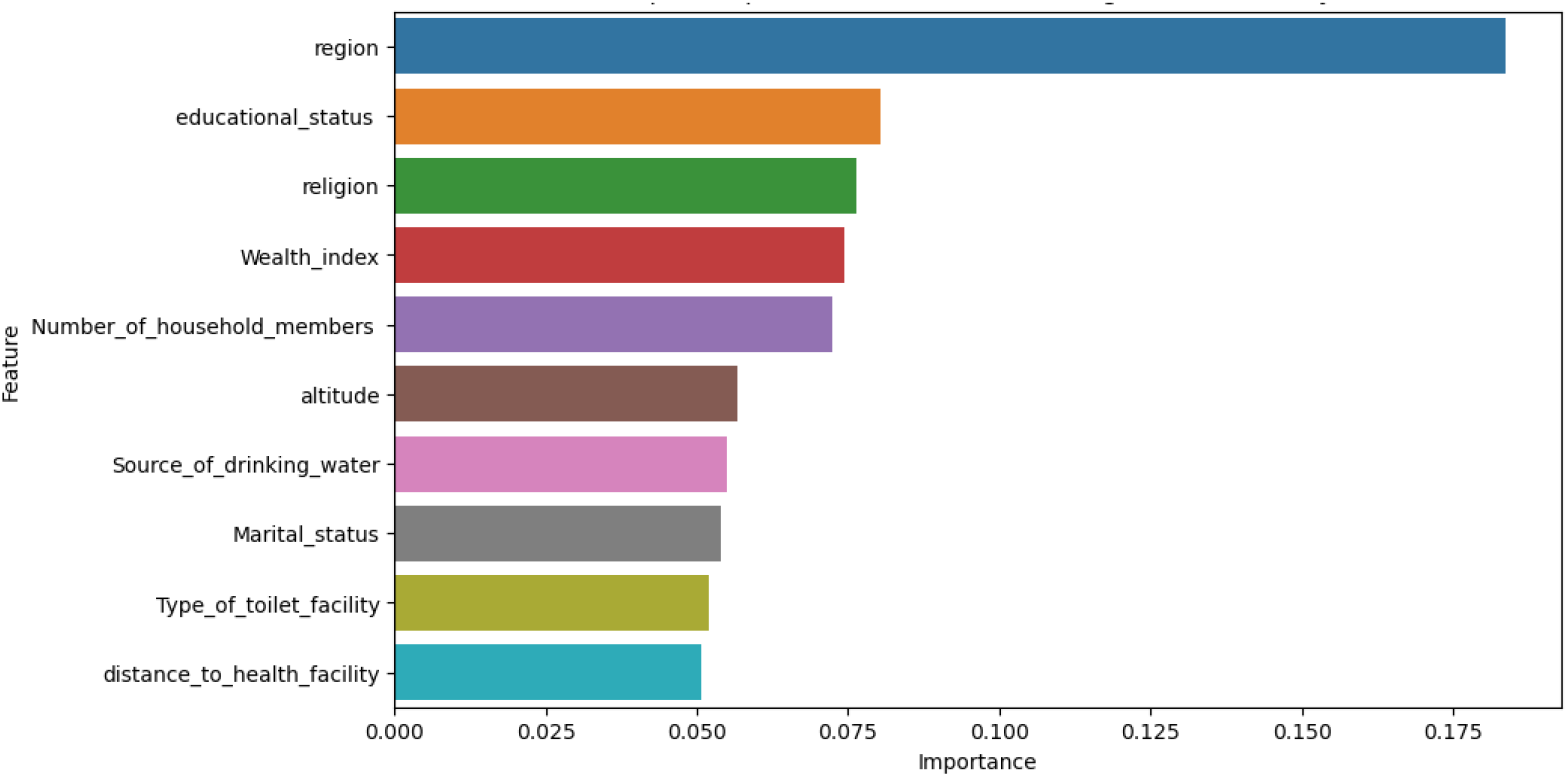
risk factor with feature importance

**Educational status** also plays a critical role, with girls having lower education levels being more vulnerable to anemia. Lack of education often correlates with limited knowledge about proper nutrition and health practices, leading to poor dietary habits that can contribute to iron deficiency. Alongside education, the **wealth index** is a significant determinant, with girls from wealthier households being at lower risk of anemia due to better access to nutritious food, healthcare, and sanitary living conditions. In contrast, girls from poorer households may face challenges in affording essential nutrients and medical care.

The **number of household members** also emerged as an important factor, suggesting that larger households may struggle with resource distribution, which could lead to poorer nutrition and increased anemia risk. The **source of drinking water** is another crucial factor, with access to clean and safe water directly influencing health outcomes. Contaminated water sources increase the risk of gastrointestinal diseases, which can worsen anemia by impairing nutrient absorption.

Cultural and social factors such as **religion** and **marital status** further shape anemia risks. Religious dietary restrictions might limit the intake of iron-rich foods, while early marriage can lead to reproductive health risks and nutritional deficiencies, which increase anemia susceptibility. Additionally, the **type of toilet facility** and **distance to health facility** highlight the importance of sanitation and healthcare access. Poor sanitation facilities and long distances to healthcare centers exacerbate anemia risk by increasing exposure to infections and limiting timely medical intervention.

In summary, the key risk factors for anemia among adolescent girls in Ethiopia are multifaceted, encompassing socio-economic, environmental, cultural, and health system-related influences. Addressing these factors requires a comprehensive approach that includes improving education, enhancing healthcare access, promoting better sanitation, and addressing socio-economic inequalities. Interventions targeting these areas could help reduce anemia prevalence and improve overall health outcomes for adolescent girls in Ethiopia.

## Conclusion

In conclusion, the evaluation of machine learning models for predicting anemia risk in adolescent girls in Ethiopia, along with the identification of key risk factors, provides a comprehensive understanding of the issue. The most risk factors are region, educational status, wealth index, and household size underscore the complex interplay of socio-economic, environmental, and cultural influences on anemia. Geographic location, access to resources, educational attainment, and socio-economic status significantly contribute to anemia risk, with factors like water source, sanitation, and healthcare access further compounding the problem.

When evaluating the machine learning models, **Random Forest** and **Extra Trees** emerge as the most effective for this classification task. These models demonstrate the highest accuracy (82.51% and 82.41%, respectively) and excellent ROC AUC scores (94.87% and 94.48%), reflecting their strong performance in distinguishing between different anemia severity levels. Their ability to handle complex interactions between risk factors makes them particularly suitable for this problem. **CatBoost** also performs robustly with a competitive ROC AUC of 93.08% and an accuracy of 80.99%, providing a solid alternative to the top two models. On the other hand, **XGBoost** and **AdaBoost** show slightly lower accuracy and ROC AUC scores, suggesting they may require further optimization to match the performance of the leading models.

Overall, **Random Forest** and **Extra Trees** are the most suitable models for predicting anemia risk in adolescent girls in Ethiopia, given their high accuracy, strong ROC AUC scores, and balanced performance across other metrics. These models, along with insights into key risk factors, can inform targeted interventions to reduce anemia prevalence, improve healthcare access, and address socio-economic disparities in Ethiopia.

## Declarations

### Author contributions

misganaw conceived and designed the study, participated in data analysis, wrote the report, finished the model refinements, carried out a deep analysis of the experiment results, drafted and revised the initial manuscript, and revised the manuscript; eyob designed the study, managed the quality and progress of the whole study, and revised the manuscript;; all authors read and approved the final manuscript.

### Data availability

http://dhsprogram.com.

## Acknowledgements

We would like to acknowledge the Ethiopian central statistics for providing us with the data with a data set description.

## Ethics declaration

not applicable because of we have used public available data by requesting to use the data

## Consent to Participate declaration

not applicable

## Reference

[1] G. A. Stevens et al., “National, regional, and global estimates of anaemia by severity in women and children for 2000–19: a pooled analysis of population-representative data,” Lancet Glob Health, vol. 10, no. 5, pp. e627–e639, 2022.

[2] S. Safiri et al., “Burden of anemia and its underlying causes in 204 countries and territories, 1990–2019: results from the Global Burden of Disease Study 2019,” J Hematol Oncol, vol. 14, pp. 1–16, 2021.

[3] R. K. Benedict, A. Schmale, and S. Namaste, “Adolescent nutrition 2000-2017: DHS data on adolescents age 15-19.,” 2018.

[4] R. T. Regasa and J. A. Haidar, “Anemia and its determinant of in-school adolescent girls from rural Ethiopia: A school based cross-sectional study,” BMC Womens Health, vol. 19, no. 1, Jul. 2019, doi: 10.1186/s12905-019-0791-5.

[5] K. A. Gonete, A. Tariku, S. D. Wami, and T. Derso, “Prevalence and associated factors of anemia among adolescent girls attending high schools in Dembia District, Northwest Ethiopia, 2017,” Archives of Public Health, vol. 76, no. 1, Dec. 2018, doi: 10.1186/s13690-018-0324-y.

[6] S. H. Tesfaye, B. T. Seboka, and D. Sisay, “Application of machine learning methods for predicting childhood anaemia: Analysis of Ethiopian Demographic Health Survey of 2016,” PLoS One, vol. 19, no. 4, p. e0300172, 2024.

[7] B. E. Dejene, T. M. Abuhay, and D. S. Bogale, “Predicting the level of anemia among Ethiopian pregnant women using homogeneous ensemble machine learning algorithm,” BMC Med Inform Decis Mak, vol. 22, no. 1, Dec. 2022, doi: 10.1186/s12911-022-01992-6.

[8] B. Tsegaye Negash and M. Ayalew, “Trend and factors associated with anemia among women reproductive age in Ethiopia: A multivariate decomposition analysis of Ethiopian Demographic and Health Survey,” PLoS One, vol. 18, no. 1, p. e0280679, 2023.

[9] K. Fentie, T. Wakayo, and G. Gizaw, “Prevalence of Anemia and Associated Factors among Secondary School Adolescent Girls in Jimma Town, Oromia Regional State, Southwest Ethiopia,” Anemia, vol. 2020, 2020, doi: 10.1155/2020/5043646.

[10] M. G. Worku, G. A. Tesema, and A. B. Teshale, “Prevalence and determinants of anemia among young (15–24 years) women in Ethiopia: A multilevel analysis of the 2016 Ethiopian demographic and health survey data,” PLoS One, vol. 15, no. 10, p. e0241342, 2020.

[11] M. G. Worku, G. A. Tesema, and A. B. Teshale, “Prevalence and determinants of anemia among young (15–24 years) women in Ethiopia: A multilevel analysis of the 2016 Ethiopian demographic and health survey data,” PLoS One, vol. 15, no. 10, p. e0241342, 2020.

[12] R. T. Regasa and J. A. Haidar, “Anemia and its determinant of in-school adolescent girls from rural Ethiopia: a school based cross-sectional study,” BMC Womens Health, vol. 19, pp. 1–7, 2019.

[13] S. H. Gebreyesus, B. S. Endris, G. T. Beyene, A. M. Farah, F. Elias, and H. N. Bekele, “Anaemia among adolescent girls in three districts in Ethiopia,” BMC Public Health, vol. 19, pp. 1–11, 2019.

[14] K. A. Gonete, A. Tariku, S. D. Wami, and T. Derso, “Prevalence and associated factors of anemia among adolescent girls attending high schools in Dembia District, Northwest Ethiopia, 2017,” Archives of Public Health, vol. 76, pp. 1–9, 2018.

[15] S. D. Habtegiorgis et al., “Prevalence and associated factors of anemia among adolescent girls in Ethiopia: A systematic review and meta-analysis,” PLoS One, vol. 17, no. 3, p. e0264063, 2022.

[16] B. Endalamaw, T. M. Abuhay, and D. Shibabaw, “Predicting the Level of Anemia among Ethiopian Pregnant Women using Homogeneous Ensemble Machine Learning Algorithm,” in Proceeding of the 2 nd Deep Learning Indaba-X Ethiopia Conference 2021, 2022.

[17] B. Kitaw, C. Asefa, and F. Legese, “Leveraging machine learning models for anemia severity detection among pregnant women following ANC: Ethiopian context,” BMC Public Health, vol. 24, p. 3500, 2024.

[18] B. E. Dejene, T. M. Abuhay, and D. S. Bogale, “Predicting the level of anemia among Ethiopian pregnant women using homogeneous ensemble machine learning algorithm,” BMC Med Inform Decis Mak, vol. 22, no. 1, p. 247, 2022.

[19] S. H. Tesfaye, B. T. Seboka, and D. Sisay, “Application of machine learning methods for predicting childhood anaemia: Analysis of Ethiopian Demographic Health Survey of 2016,” PLoS One, vol. 19, no. 4, p. e0300172, 2024.

[20] A. Kebede Kassaw, A. Yimer, W. Abey, T. L. Molla, and A. B. Zemariam, “The application of machine learning approaches to determine the predictors of anemia among under five children in Ethiopia,” Sci Rep, vol. 13, no. 1, p. 22919, 2023.

[21] A. B. Zemariam et al., “Employing advanced supervised machine learning approaches for predicting micronutrient intake status among children aged 6–23 months in Ethiopia,” Front Nutr, vol. 11, p. 1397399, 2024.

[22] A. B. Zemariam et al., “Employing supervised machine learning algorithms for classification and prediction of anemia among youth girls in Ethiopia,” Sci Rep, vol. 14, no. 1, p. 9080, 2024.

